# 36-h Total Sleep Deprivation Induced Aberrant Insula Sub-regional Functional Connectivity

**DOI:** 10.1101/2023.06.09.23291196

**Authors:** Xiangling Chen, Kaiming Zhang, Shiyu Lei, Hai Yang, Yue Zheng, Xuemei Wu, Xinuo Ma, Xiechuan Weng

**Affiliations:** Department of Public Health, Chengdu Medical College, Sichuan, 610500, China; Department of Clinical Medicine, Chengdu Medical College, Sichuan, 610500, China; Department of Pharmacology, Chengdu Medical College, Sichuan, 610500, China; Department of Neuroscience, Beijing Institute of Basic Medical Sciences, Beijing, 100850, China

**Keywords:** rs-fMRI, insular subregions, PVT, sleep deprivation, functional connectivity

## Abstract

Total sleep deprivation (TSD) induces aberrant insula functional connectivity (FC). The insula comprises at least three functionally distinct subregions: anterior dorsal (dAI), anterior ventral (vAI), and posterior insula (PI). Biased attention toward the anterior insula (AI) has limited our complete understanding of the TSD impact. We aimed to investigate TSD-induced functional connectivity and lateralization across the insula subregions. A total of 54 healthy young men completed 36-h TSD. Two sessions of psychomotor vigilance task (PVT) and 3T resting-state functional magnetic resonance imaging (rs-fMRI) scanning were carried out. A seed-based FC was conducted using bilateral insula subregions. Impaired vigilance, altered functional connectivity and lateralization were observed after TSD. The AI showed enhanced connectivity with the cerebellum, middle frontal gyrus, putamen, and postcentral gyrus but decreased connectivity with the temporal lobes, angular gyrus, calcarine sulcus, anterior cingulum, and medial orbitofrontal gyrus. The PI had increased connectivity with the middle frontal, inferior temporal, and inferior parietal gyrus but showed an anti-correlation with the middle temporal regions, posterior cingulum, and angular gyrus. All the seeds showed ipsilateral connections with specific brain regions, excluding the cerebellum. Both the vAI and PI displayed FC with the insula. The correlation analysis between PVT and brain signal changes did not survive Bonferroni correction. This study provided information about potential functional asymmetries of insula subregions caused by 36-h TSD. These findings provided new insights into the neural mechanisms of inter-hemispheric communication and coordination, which is essential for understanding the overall brain function.

## Introduction

Sustained attention refers to the ability to stay focused in front of a stimulus within a specific time frame. It is a form of higher-order cognitive function. Several findings have validated that sustained attention is susceptible to sleep deficiency [1–3]. The brain regions closely interrelated to cognition modulation and vulnerable to sleep loss include frontal and parietal areas (cognitive control), secondary processing cortices (for example, pre/supplementary motor area), and thalamic regions [4]. Sleep deprivation disrupts the normal functioning of highly organized brain networks. One night of sleep loss impairs the functional integrity of the default mode network (DMN) [5] and affects internal information processing that is basic for human activities: language [6], perception [7, 8], emotion [9], and memory [10, 11]. Strong anti-correlation between DMN and attention network is observed after sleep deprivation [12].

Insula is among one of the major hubs that regulate sustained attention.

Previous studies have shown that after 36 h of total sleep deprivation (TSD), aberrant functional connectivity (FC) pattern was observed between the insula and anterior cingulate cortex (ACC), prefrontal, temporal, parietal, and occipital areas. The altered insula-prefrontal connection was significantly correlated with vigilant attention[13]. However, in this study, we only assessed the FC pattern of the insula as a whole. Network analysis provided a refined result by reporting enhanced anterior insula (AI) connection with ACC after 36-h TSD. A positive correlation was established between such connectivity and response time in cognitive tasks [14]. A recent study investigated the involvement of AI in TSD-impaired cognition, dividing the insula into two segments, ventral and dorsal. Using resting-state functional magnetic resonance imaging (rs-fMRI), a laterality effect was observed between bilateral dorsal anterior insula (dAI) seeds—the left dAI showed an increased connection with the right superior frontal gyrus (SFG), while the right was significantly connected to the bilateral SFG and right putamen. The left dAI-SFG FC, not the contralateral hemisphere, was statistically correlated with impaired cognitive outcomes [12].

These findings raise two questions. First, although there is evidence that the insula has at least three functionally distinct subregions (ventral, dorsal, and posterior), the posterior insula (PI) has received less attention, which has limited our understanding of its role in higher-order functions. Some studies have suggested that the PI may be involved in sustained attention [15, 16]. Additionally, it has been shown to have functional connections with other brain regions involved in attention and cognitive control [12]. However, the correlation between PI and sustained attention is yet to be elucidated. Second, whether the lengths of TSD (24 h *vs*. 36 h) generate different FC patterns is yet unknown. Previous studies have omitted the discussion about the PI, and hence, we can only compare the FC of AI after 24-h TSD and 36-h TSD. On the other hand, numerous studies have reported hemispheric asymmetry in regions regulating cognitive functions [17]. The investigation of the impact of 36-h TSD on the functional lateralization of insula subregions would provide significant insights into the neural mechanisms of inter-hemispheric communication and coordination, which is crucial for comprehending the brain’s overall function. We hypothesized that 36-h TSD exerts a laterality effect on three insular subparts, and the altered FC pattern is linked to impaired sustained attention.

## Methods

### Ethics

All participants provided written informed consent, and the study was approved by the Institutional Review Board of Beihang University(Beihang University, Beijing, China). Approval number: BM20180040.

### Sample size

We used a sample size similar to those of previous work[18].

### Design and participants

A total of 54 right-handed male volunteers (age 20–29 years, M±SD: 21±2 years) completed the 36-h TSD, including two sessions of rs-fMRI (both at 20:00) and PVT scheduled before and after TSD. Subjects had no history of psychological or neuronal impairment. No sleep problem was declared. No alcohol or caffeine intake was reported 2 days prior to the trial. All subjects were college students with a relatively fixed sleep schedule from 10:30 p.m. to 7:30 a.m.

**Figure 1.**
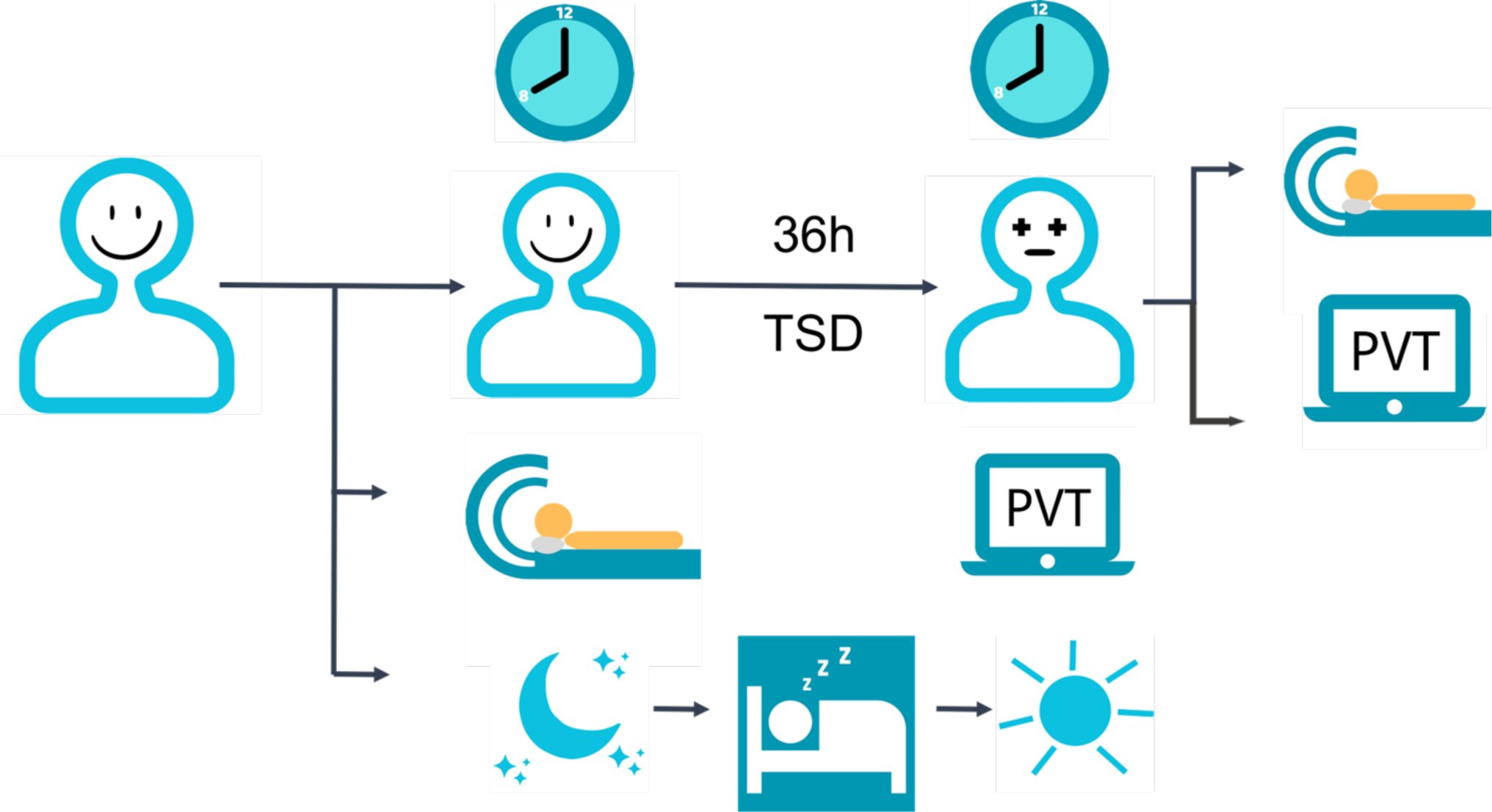
General workflow

### Vigilance measurement

The standard 10-min PVT version was used to assess the alertness level induced by 36-h TSD. Randomized inter-stimulus interval (ISI) was set to 2–10 s. The participants were instructed to focus on the laptop screen and click the mouse as soon as a visual stimulus (an Arabic number that keeps turning bigger if there is no reaction) appeared. PVT outcomes were calculated based on the subjects’ performance between response to the last stimulus and the appearance of the next one. Response time (RT) > 500 ms was defined as “Minor Lapses.” The shorter the RT, the higher the level of vigilant attention. However, early response, i.e., clicking the mouse before the appearance of the next stimulus, was considered “False Starts (FS).” In addition, an “FS” warning would be displayed on the screen. The most-commonly used outcomes included for analysis were as follows: (1) number of lapses (RTs ≥ 500 ms); (2) number of false starts; (3) number of lapses and false starts; (4) mean RT; (5) mean 1/RT (also known as reciprocal RT or response speed) [19], [20]; (6) standard deviation RT (StdDev RT); (7) median RT; (8) minimum RT; (9) maximum RT; (10) fastest 10% RT; (11) slowest 10% RT; (12) speed.

### Image acquisition

Brain images were acquired on a 3-T GE scanner (Discovery MR750w 3.0T, General Electric Medical Systems, Wisconsin, USA). High-resolution T_1_-weighted structural images of the whole brain were acquired with a 3D MPRAGE sequence (TR = 2 s, TE = 30 ms, FA =12°, FOV = 256 × 256 mm^2^, voxel size = 1 × 1 × 1 mm^3^, 192 slices, and matrix = 64 × 64). 8-min resting-state blood-oxygen-level-dependent (BOLD) scans were collected with 90° flip angle, TR/TE = 2000/30 ms, FOV = 256 × 256 mm^2^, matrix = 64 × 64, and 3-mm slice thickness with 1-mm slice gap. The subjects were instructed to remain awake with their eyes closed. No participant had head motion exceeding 2.0 mm of maximal displacement and 2.0 of maximal rotation in any direction.

### rs-fMRI analysis

Data were preprocessed using the Statistical Parametric Mapping (SPM) 12 software (http://www.fil.ion.ucl.ac.uk/spm/)under MATLAB and DPABI toolbox [21]. The first ten volumes from each resting dataset were removed to allow magnetization to a steady state. The preprocessing pipelines included slice timing and realignment for head motion correction. Functional data were coregistered to high-resolution anatomical images, which were, in turn, normalized to Montreal Neurological Institute (MNI) space at 3 × 3 × 3 mm^3^. Nuisance covariates (Friston 24 head motion parameters, grey matter signals, white matter signals, and cerebrospinal fluid signals) were regressed. The data were further spatially smoothed using 6-mm full-width at half maximum (FWHM) Gaussian kernel. To reduce the effect of low-frequency drifts and high-frequency noises, a band-pass temporal filtering at 0.01–0.1 Hz was applied. Then, a seed-to-voxel approach was conducted. The pre-defined regions of interest (ROIs) were placed in bilateral insulae and further separated into three functional regions based on the anatomical features: dorsal anterior insula (dAI), ventral anterior insula (vAI), and posterior insula (PI) [22]. To investigate the lateralization after 36-h TSD, six seeds were used to calculate the correlation coefficient with the whole brain voxels using paired-t tests (left vs. right). The connectivity maps were then *z*-standardized and introduced to group-level analysis using a paired *t*-test. At the group level, correction for multiple comparisons was conducted using Gaussian random field theory to threshold significant voxels at P < 0.01 and clusters at P < 0.01. Subsequently, the correlation coefficients of the peak connectivity voxels(cluster size>400) for the six insular subparts were extracted and reported using MNI labels. Finally, these values were used to investigate potential correlations with PVT outcome variables.

## Results

### Sleep-deprived PVT performance

The level of vigilance attention decreased significantly after 36 h of sustained wakefulness, as indicated by a significant number of lapses, false starts, and the sum of the two items. A longer response time, slower speed, and higher lapse probability were reported. After multiple comparisons correction, it was found that all are P < 0.05 except for minimum RT.

**Table 1.**
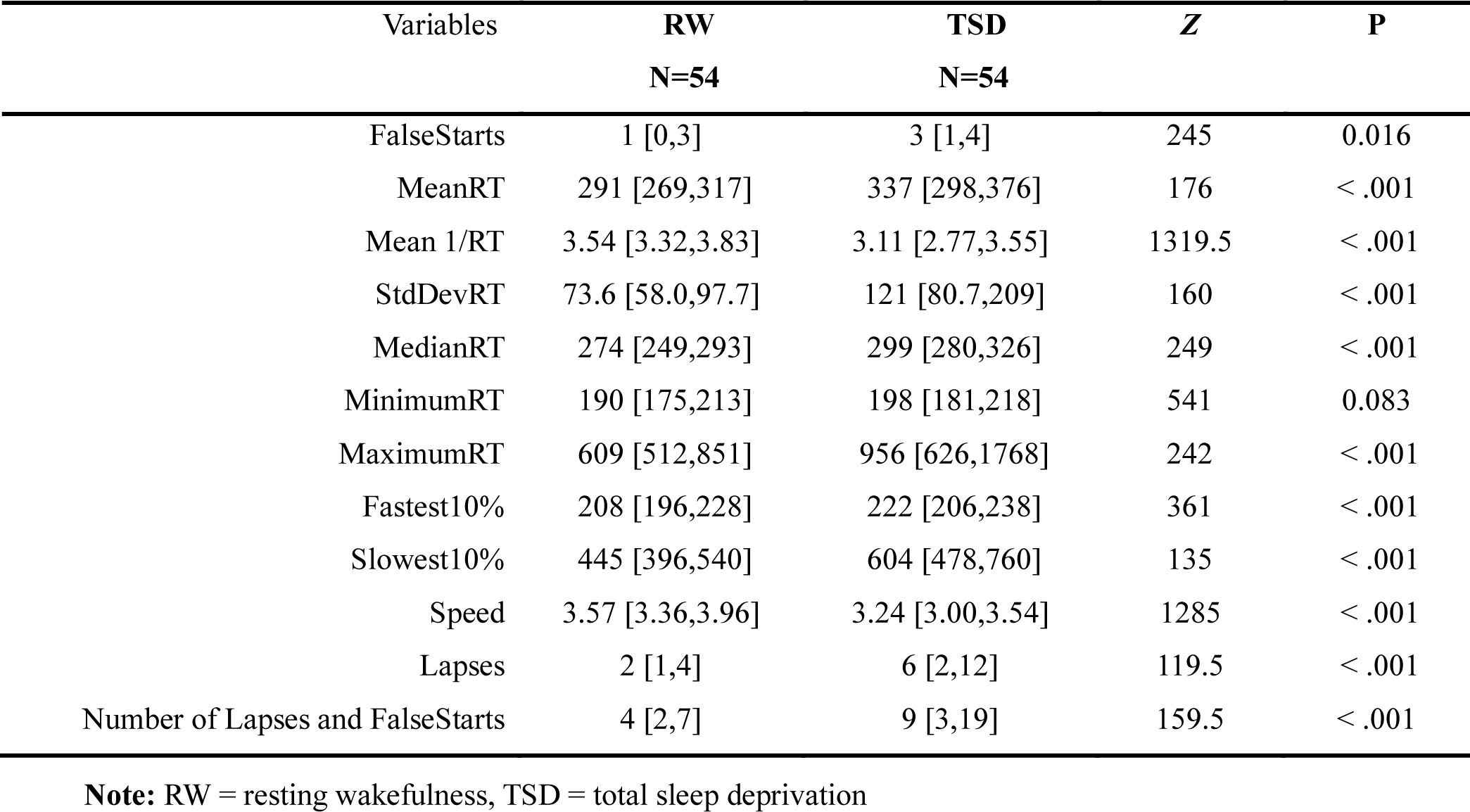
Comparisons of PVT outcomes after 36-h TSD

| Variables | RW<br>N=54 | TSD<br>N=54 | Z | P |
| --- | --- | --- | --- | --- |
| FalseStarts | 1 [0,3] | 3 [1,4] | 245 | 0.016 |
| MeanRT | 291 [269,317] | 337 [298,376] | 176 | < .001 |
| Mean 1/RT | 3.54 [3.32,3.83] | 3.11 [2.77,3.55] | 1319.5 | < .001 |
| StdDevRT | 73.6 [58.0,97.7] | 121 [80.7,209] | 160 | < .001 |
| MedianRT | 274 [249,293] | 299 [280,326] | 249 | < .001 |
| MinimumRT | 190 [175,213] | 198 [181,218] | 541 | 0.083 |
| MaximumRT | 609 [512,851] | 956 [626,1768] | 242 | < .001 |
| Fastest10% | 208 [196,228] | 222 [206,238] | 361 | < .001 |
| Slowest10% | 445 [396,540] | 604 [478,760] | 135 | < .001 |
| Speed | 3.57 [3.36,3.96] | 3.24 [3.00,3.54] | 1285 | < .001 |
| Lapses | 2 [1,4] | 6 [2,12] | 119.5 | < .001 |
| Number of Lapses and FalseStarts | 4 [2,7] | 9 [3,19] | 159.5 | < .001 |

**Note:** RW = resting wakefulness, TSD = total sleep deprivation

### RW vs. TSD: seed-to-voxel analysis

Compared to baseline, the right dAI showed increased FC with the cerebellum_R but decreased FC with middle temporal gyrus(MTG),_R superior temporal gyrus(STG)_R, and angular gyrus(AG)_L. (**Figure 2a**).

**Figure 2.**
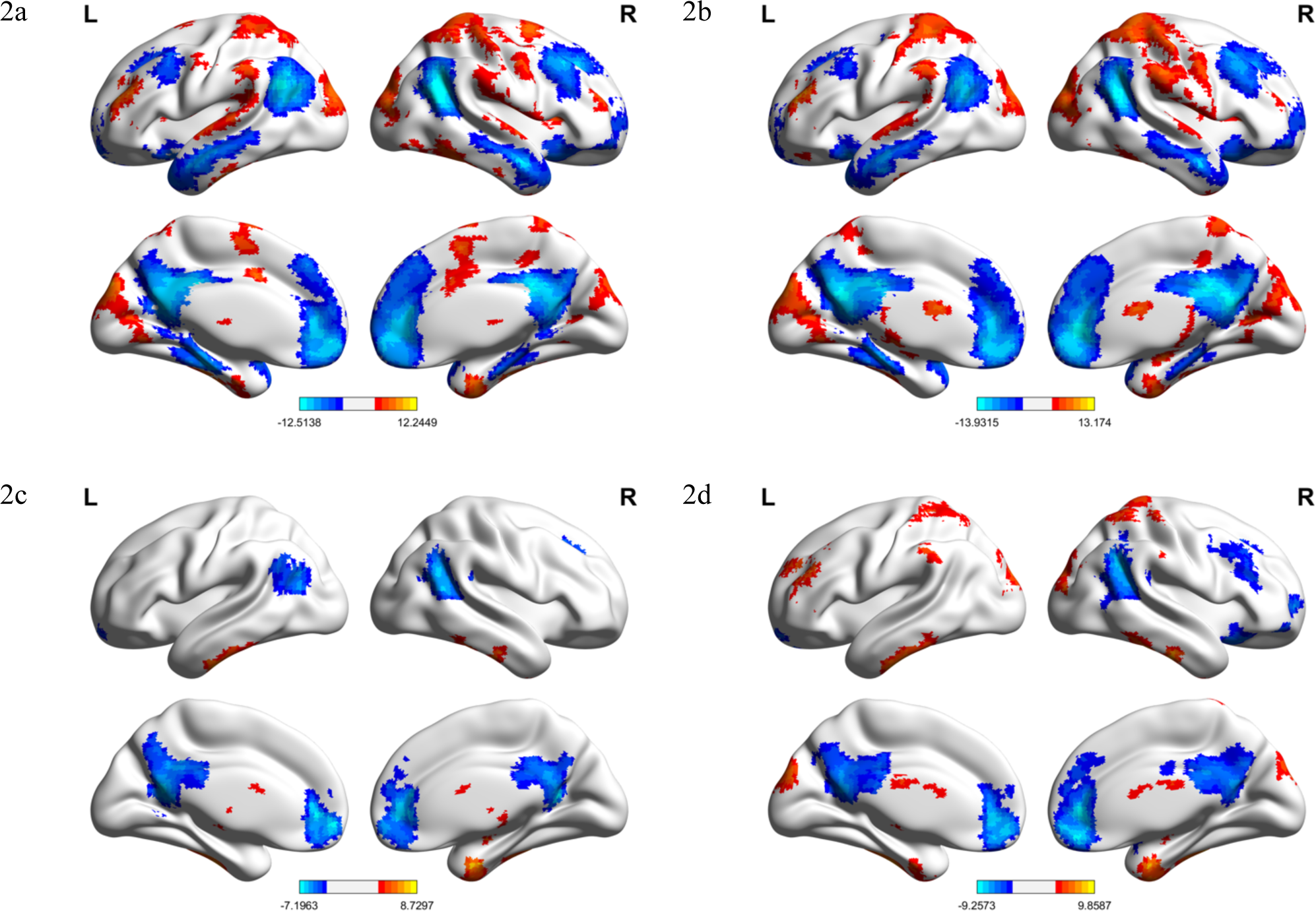
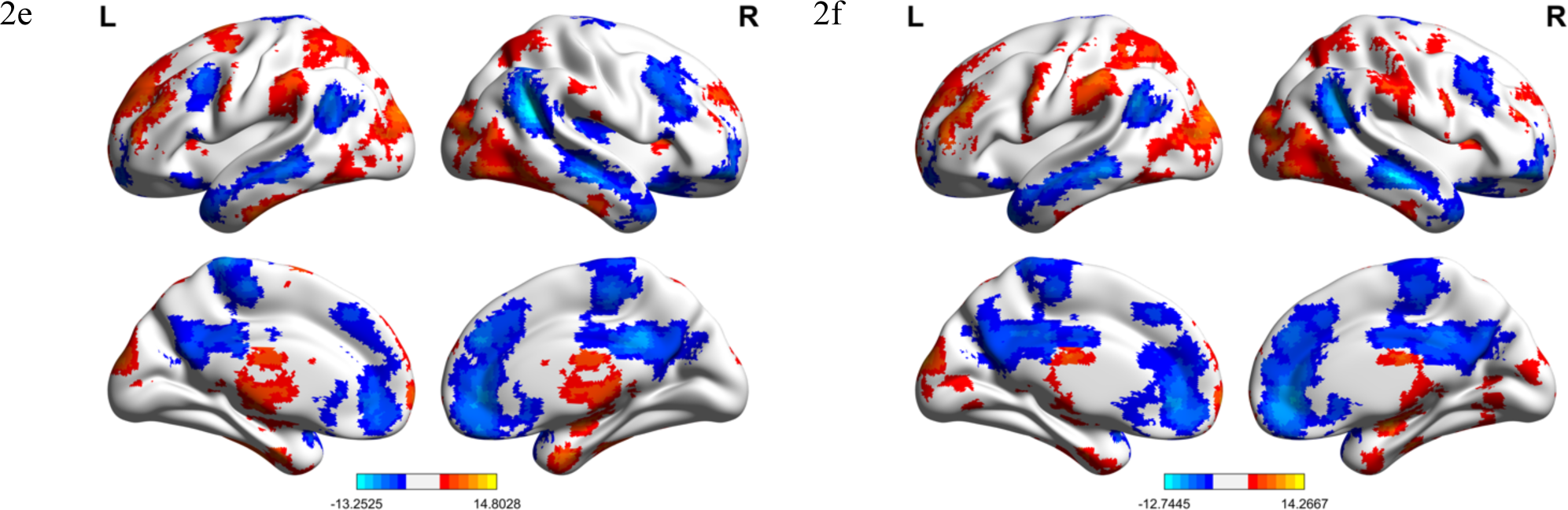
Functional connectivity patterns of bilateral insular subregions induced by 36-h TSD. Cold colors indicate decreased connectivity after 36-h TSD, and warm colors represent increased connection after 36-h TSD.

**Table 2.**
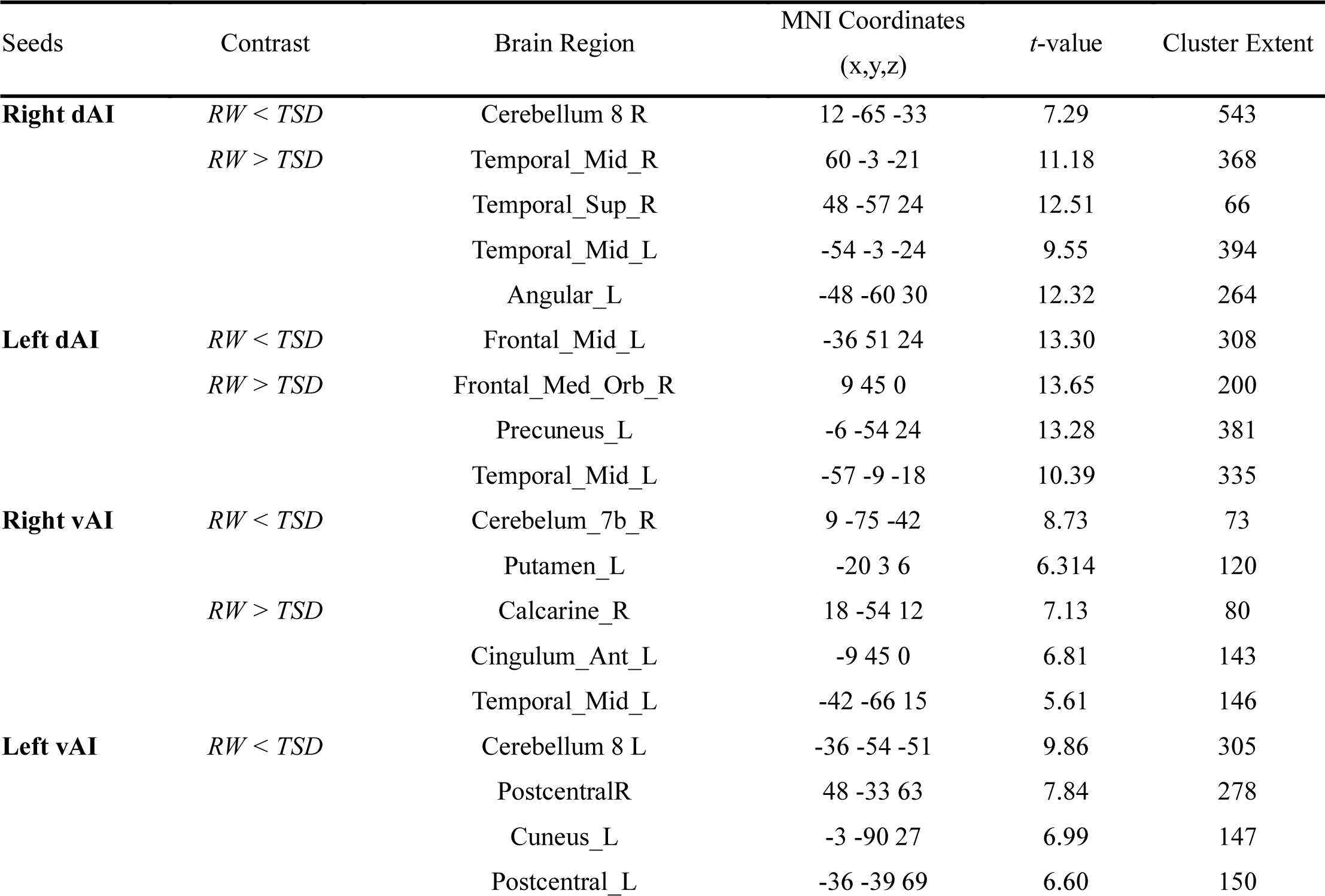
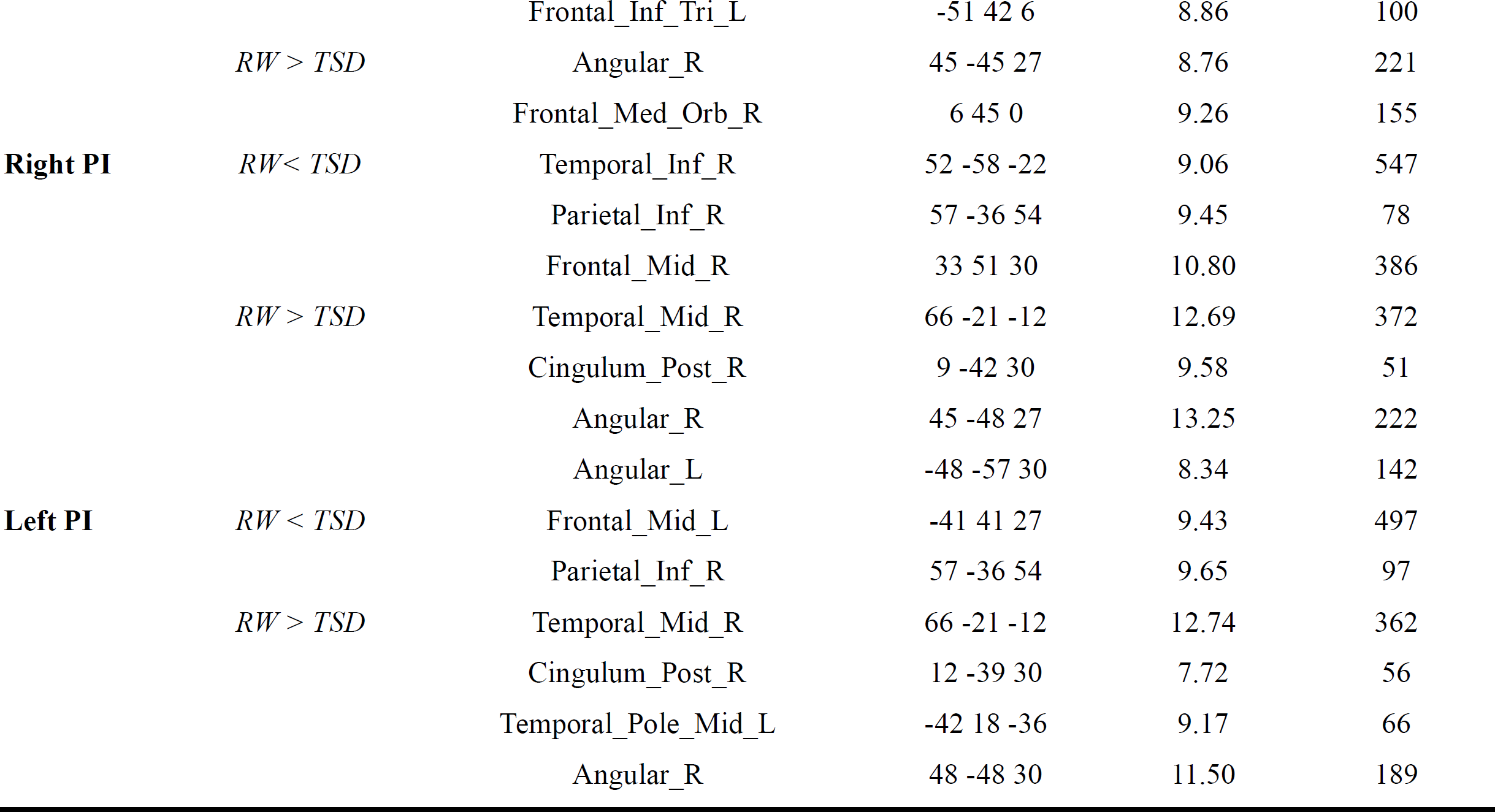
Seed-based FC after 36-h TSD

After 36h-TSD, the left dAI had enhanced connections with middle frontal gyrus(MFG)_L, while weakened FC with frontal medial orbital gyrus(MOG)_R, precuneus_L, and MTG_L(**Figure 2b**).

The right vAI had elevated FC with the cerebellum_R and putamen_L, but decreased FC with calcarine sulcus_R, anterior cingulum(AC)_L, and MTG_L(**Figure 2c**).

The left vAI displayed increased FC with the cerebellum_L, postcentral gyrus(PoCG), cuneus_L and inferior frontal gyrus(IFG)_L(**Figure 2d**).

The right PI had enhanced FC with inferior temporal gyrus(ITG)_R, inferior parietal gyrus(IPG)_R, and MFG _R, but decreased FC with regions including MTG_R, posterior cingulum(PC)_R and bilateral AG post-TSD(**Figure 2e**).

The left PI displayed stronger connections with right ITG, IPG and MFG, while weaker connections with right MTG, PC, AG and left middle temporal pole(**Figure 2f**).

### Analysis of laterality effects

This study suggested a lateralized effect on insula sub-regional FC by comparing the FC of seeds in both the left and right hemispheres. The results showed ipsilateral connections with specific brain regions (excluding the cerebellum) for all seeds. Additionally, the vAI and PI showed FC with the insula (**Table 3 and Figure 3**).

**Figure 3.**
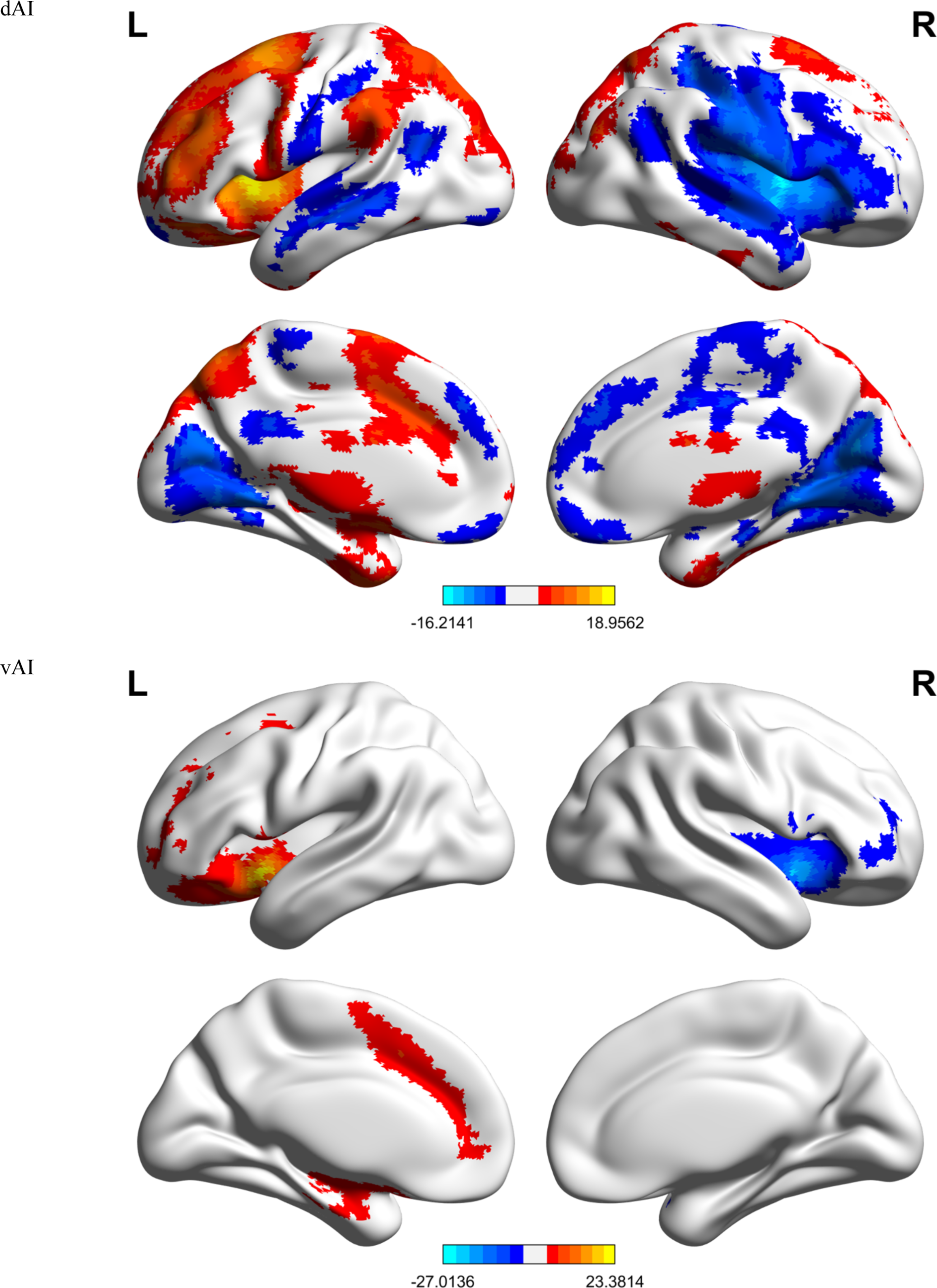
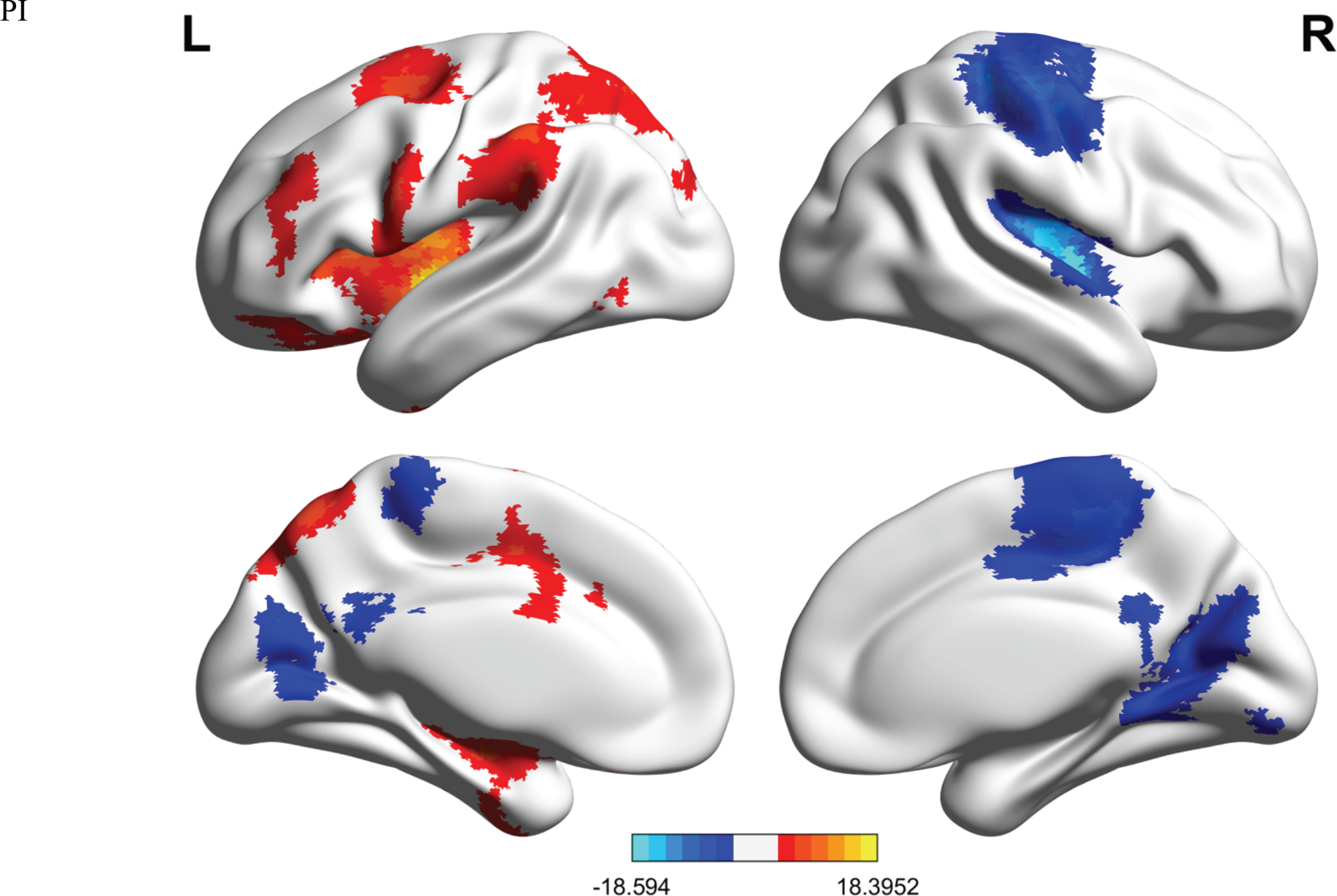
TSD-induced FC asymmetry. Cold colors indicate stronger FC in the left compared to the right hemisphere.

**Table 3.** TSD-induced laterality effect

| Seeds | Contrast | Brain Region | MNI Coordinates | *t*-value | Cluster Extent |
| --- | --- | --- | --- | --- | --- |
|  |  |  | (x,y,z) |  |  |
| **dAI** | L > R | Frontal_Mid_L | -28 40 21 | 5.54 | 1009 |
|  |  | Cerebelum_Crus1_R | 40 -58 -39 | 9.05 | 152 |
|  | L < R | Temporal_Sup_R | -51 -4 0 | 7.76 | 672 |
|  |  | Calcarine_R | 3 -72 18 | 10.73 | 332 |
| **vAI** | L > R | Insula_L | -36 9 -9 | 23.38 | 196 |
|  | L < R | Insula_R | 39 12 -9 | 27.01 | 347 |
| **PI** | L > R | Insula_L | -38 -9 -3 | 17.79 | 425 |
|  |  | Precentral_L | 29 -11 51 | 5.55 | 194 |
|  | L < R | Insula_R | 39 -9 0 | 18.60 | 187 |
|  |  | Cuneus_L | 0 -72 24 | 7.20 | 123 |

### Associations between altered insular connectivity and poor PVT performance

In order to further elucidate brain-behavior correlations between insula subregions and impaired vigilant attention, Spearman’s rank correlation analysis was carried out for subparts and PVT outcome variables. No correlation survived after Bonferroni’s correction(**Supplementary Fig.1**).

## Discussion

Unraveling the mechanistic of declined sustained attention is a pivotal aspect of understanding human brain functions. Our study aimed to investigate the impact of 36 h of TSD on the FC of bilateral insular subregions and to examine the correlation between these changes and any impaired psychomotor vigilance. Additionally, we also determined any laterality effects on the FC of these ROIs. The data revealed that TSD prolonged the response time and speed. The AI had increased FC with the cerebellum, MFG, putamen, and PoCG but decreased connections with superior and middle temporal gyri, AG, PFC, precuneus, calcarine sulcus, and AC. The PI had strong connections with the MFG, ITG, and IP but showed an anti-correlation with the MTG, PC, and AG.

The cerebellum is a motor center and is also involved in cognitive processes, such as attention, language, and emotional regulation [12]. Lesion studies and neuroimaging observations have repeatedly observed cerebellar contribution to cognitions through the integration with regions, such as the frontal and prefrontal cortices [23–25]. The PFC, with its dense connections, is involved in a range of higher-order cognitive functions and sensory motor control, making it the site of the highest level of neural integration [26, 27]. The cortical thickness of the insula and PFC is associated with aggressive behaviors [28]. This finding is relevant to the functional integration between the cerebellum and frontal cortex, which could be explained by the cerebellum’s ability to create internal simulations at the cognitive level, modulating non-motor processing [29, 30].

The AI, MFG, temporo-parietal junction (IPC), AC, and intraparietal sulcus belong to large brain networks, including the dorsal (DAN) and ventral attention networks (VAN) [31, 32]. These hubs have been implicated in exerting top-down executive controls on various aspects [33]. The MFG is a convergence site for the DAN and VAN, where it reorients attention from exogenous to endogenous control [34, 35]. It is also a key component of the dorsal lateral PFC (DLPFC) that is involved in attention control [36–38]. Some studies revealed that the DLPFC is strongly connected to frontal cortices that are preferentially involved in motor control [30]. FMRI studies frequently reported coactivation between the DLPFC and cerebellum during cognitive tasks [39–41]. Moreover, the application of repetitive transcranial magnetic stimulation (rTMS) and transcranial direct current stimulation (tDCS) to the DLPFC resulted in core symptom (emotion and cognition) improvement on major depression disorder (MDD) [42, 43]. Neuroimaging studies revealed that aberrant regional signals within the DLPFC may be potential biomarkers for differentiating the treatment resistant depression (TRD) from non-TRD [44, 45]. Taken together, these results suggested that MFG_L is sensitive to regional and brain level FC. The DLPFC may be the neural underpinning of emotional and attentional impairment caused by sleep deprivation. Moreover, the AC is involved in various aspects of executive function and decision making. Altered cingulum bundle microstructure has been linked to cognitive control in psychiatric conditions and has been widely reported in the literature [46–48]. Similarly, the IPC represents a complex behavioral and cognitive repertoire, including sustained attention [49], somatosensory-motor integration [50, 51], and execution [52] due to its location (within the inferior parietal lobule and the overlapping temporoparietal junction). Desmurget and Sirigu proposed a two-step process for motor intention triggered by goal-directed actions, in which the IPC controls the initial “wanting to move” intention and specifies a general goal before motor planning [53]. MCI patients showed significantly different deoxygenated-hemoglobin (HbR) concentrations within the inferior parietal regions during the task, indicating a potential for IPC in the initial assessment of MCI [54].

Anderson et al. demonstrated that the connectivity patterns in the insula and putamen networks are correlated with self-rated cognitive fatigue (CF), suggesting its potential involvement in the subjective experience of CF [55]. Putamen abnormalities in the structure and function are potential biomarkers for individuals at high risk for MDD [56]. Complex motor stereotypes (CMS) may be potentially linked to motor and cognitive deficits [57]. Another study showed that children with CMS have a reduced volume size within the putamen [58]. Consistent with a previous study, we observed altered interactions between insula subparts and regions, including putamen, PoCG, and DLPFC. In addition, the changes in connectivity patterns were statistically correlated with anxiety, depression, and cognitive impairment [59]. These findings suggested that the insula plays a critical role in functional integration with other brain regions for sensory integration, cognitive, and emotional processing. The dysfunction of this connectivity may contribute to various psychological disorders.

Although both the PoCG and visual cortex are involved in motor control, the visual cortex specifically plays a role in regulating visual attention [60, 61]. The cuneus and calcarine sulcus are responsible for visual information processing [62–65]. According to Raven et al., sleep induces synaptic potentiation within neurons of the primary visual cortex [66]. The strengthened communication to the cuneus but decreased to the calcarine sulcus could be interpreted as a compensatory effort in response to sleep deficiency. Such dynamic interaction patterns may be explained by a counter-TSD effort.

The precuneus and surrounding regions are considered hot spots in the brain due to their high resting metabolic rates. Thus, it could be inferred that the precuneus plays a role in the network of neural correlates responsible for self-awareness and is involved in self-referential thinking during rest [67]. On the other hand, the insula is a core hub in the executive control network. Our study showed that 36-h TSD has impaired functional coordination between the two major networks.

Ramage et al. indicated the role of MTG in subjective fatigue [68]. Luo et al. investigated the effect of sleep disorder on aberrant network connectivity in mild cognitive impairment. The study also found that the MCI-SD group had stronger connectivity between the default mode network (DMN) and left MTG [69]. Gray matter volume reductions in the MTG have been associated with functional deficits in cognitive processing [70], and hypometabolism of the MTG is a sign of subjective cognitive decline in preclinical stages of neurological disorders [71]. The existing evidence also suggested that the FC between the STG_R and insula_R may be related to social perception and emotional processing [72]. Weakened signal transmission between the insula_R and STG in schizophrenia patients may be linked to deficits in generating and delivering a fluent speech, as well as to the coherence and logic of thought processes [73]. However, the role of MTG in language processing is widely accepted [74]. Although our study did not investigate the impact of TSD on language processing, it is plausible that cognitive functions related to language generation and processing could be affected by sleep disturbances, owing to the impact of sleep on cognitive function.

According to the literature, the TSD-induced enhanced FC between the PI and ITG may be related to visual processing [75, 76]. On the other hand, AG is an essential part of the parietal system. Previous studies have documented the AG’s involvement in cognitive processing, including attention and awareness, as well as its role in visuospatial processing [77–80]. The current study revealed a significant decrease in AG signals after sustained wakefulness, which is consistent with previous studies on children with obstructive sleep apnea. This finding suggested a potential link between reduced AG activity and impaired cognition [81].

Delano-Wood et al. demonstrated that the disruption of white matter integrity in the PC was linked to memory deficits and verbal memory performance in individuals with mild cognitive impairment (MCI). The study also suggested that measuring posterior cingulum integrity could be a potential biomarker for cognitive decline in MCI and Alzheimer’s disease [82]. Another study investigated the correlation between mean diffusivity (MD) in the PC and disease progression in AD patients. The results showed a significant correlation between MD in the PC and disease progression, deeming it a sensitive indicator of cognitive impairment [83].

The current study revealed that 36 h of TSD can cause functional asymmetries in the insula subregions. Our findings indicated that TSD has a specific impact on the connectivity of these subregions, based on the comparison of the FC between the seeds in the left and right hemispheres, implying that TSD affects the communication between the insula and other brain regions, as well as the internal connections within the insula itself. These findings offer new insights into the neural mechanisms of inter-hemispheric communication and coordination and the brain’s processing of sensory information and modulation of emotional responses under TSD influence.

Moreover, the reported laterality effects could have potential implications on our understanding of the overall brain function. The knowledge of how TSD affects the laterality of the insula could be vital in diagnosing and treating sleep-related disorders and related diseases, such as chronic fatigue, depression, and neurodegenerative disorders. However, additional studies are warranted to validate these results and explore their potential implications.

However, the correlation between PVT performance and brain signal changes did not survive the correction for multiple comparisons, highlighting the need for further studies to fully understand the association of TSD with brain function. Also, the exclusion of female participants limits the generalization of our results.

## Conclusion

The present study aimed to investigate the impact of 36-h TSD on sustained attention, FC patterns, and functional lateralization of insula subordinate regions. rs-fMRI data revealed that AI had more extensive connections with brain regions (cerebellum, MTG, STG, AG, MFG, PFC, precuneus, putamen, visual cortex, AC, and PoCG) than PI (MFG, ITG, IPG, MTG, PC, and AG). These findings have significant implications for understanding the neural mechanisms underlying sleep deprivation and its effects on cognitive and brain functions. Future studies would explore how TSD affects other brain regions and their FC patterns. Additional investigation into the mechanisms underlying these changes in FC may shed light on potential interventions for sleep-deprived individuals. Finally, it may be beneficial to examine how sleep deprivation affects other cognitive processes beyond sustained attention.

## Declarations

### Conflict of Interest

The authors declare that the study was conducted in the absence of any commercial or financial relationships that could be construed as a potential conflict of interest.

### Author contribution

Xiangling Chen: Methodology, Software, Formal analysis, Data curation, Writing, Visualization, Figure drawing; Kaiming Zhang: Software, Data curation; Shiyu Lei: Invesigation, Validation; Hai Yang: Resources, Investigation; Yue Zheng: Investigation; Xuemei Wu: Resources; Xinuo Ma: Investigation; Xiechuan Weng: Funding acquisition, Supervision.

## Data Availability

All data produced in the present study are available upon reasonable request to the authors

## Acknowledgments

We thank the undergraduate team for conducting this trial: Yidan Wang, Dan Wang, Weihong Lai, Qingshuang Lu, Yao Deng, Jing Li, Yaru Sun, Huanlin Yang, Qi Kong, Yuzhi Ming, and Yiyang Yu.

## Funding

This study was supported by the National Key R&D Plan (2022YFC3600500), and the National Natural Science Foundation of China (No. 82073833 and 8216050478).

## Data Availability Statement

All data will be made available upon reasonable request.

## Supplementary figures

**S1. Spearman.**
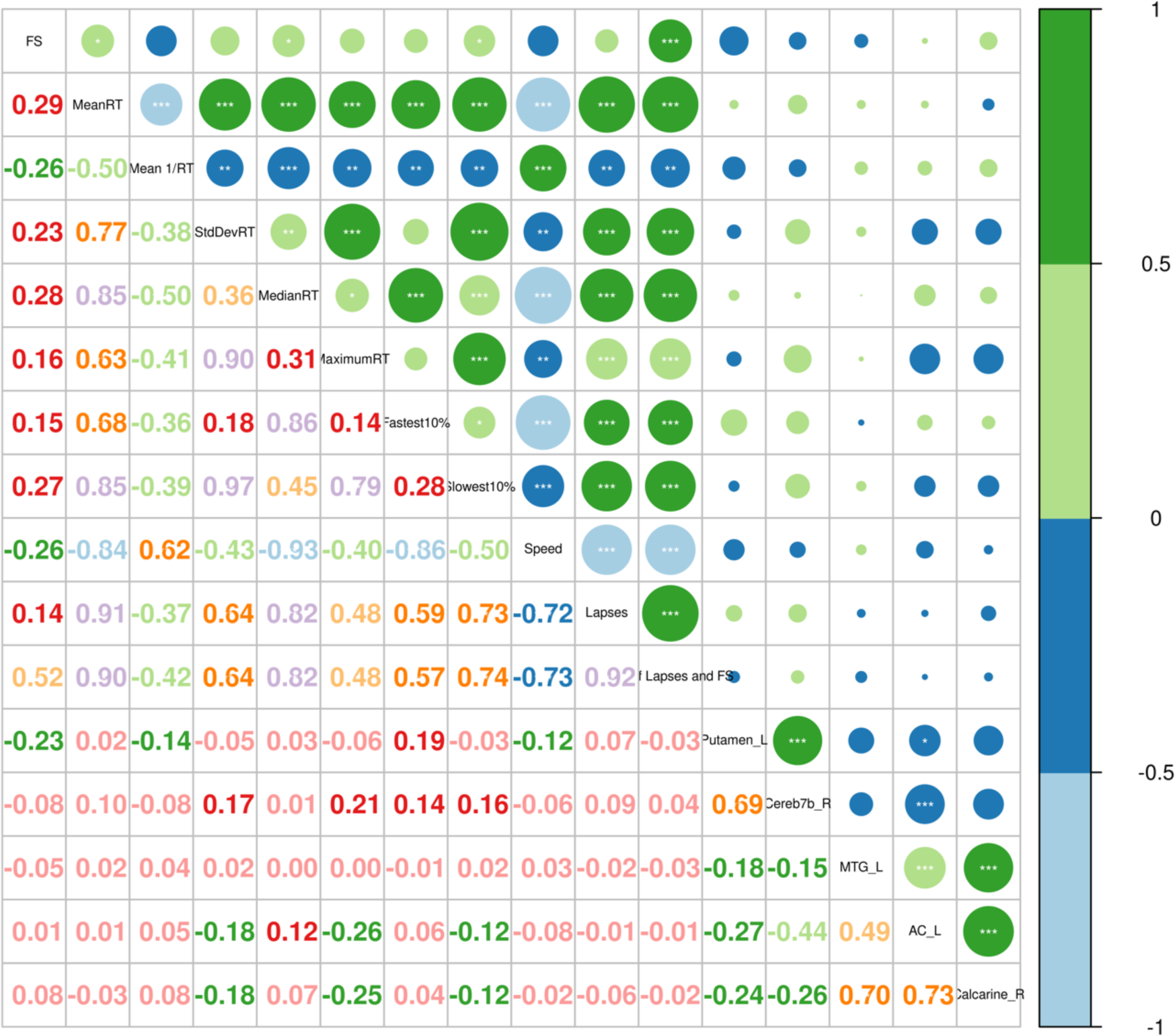
Correlation analysis between vAI_R & PVT

**S2. Spearman.**
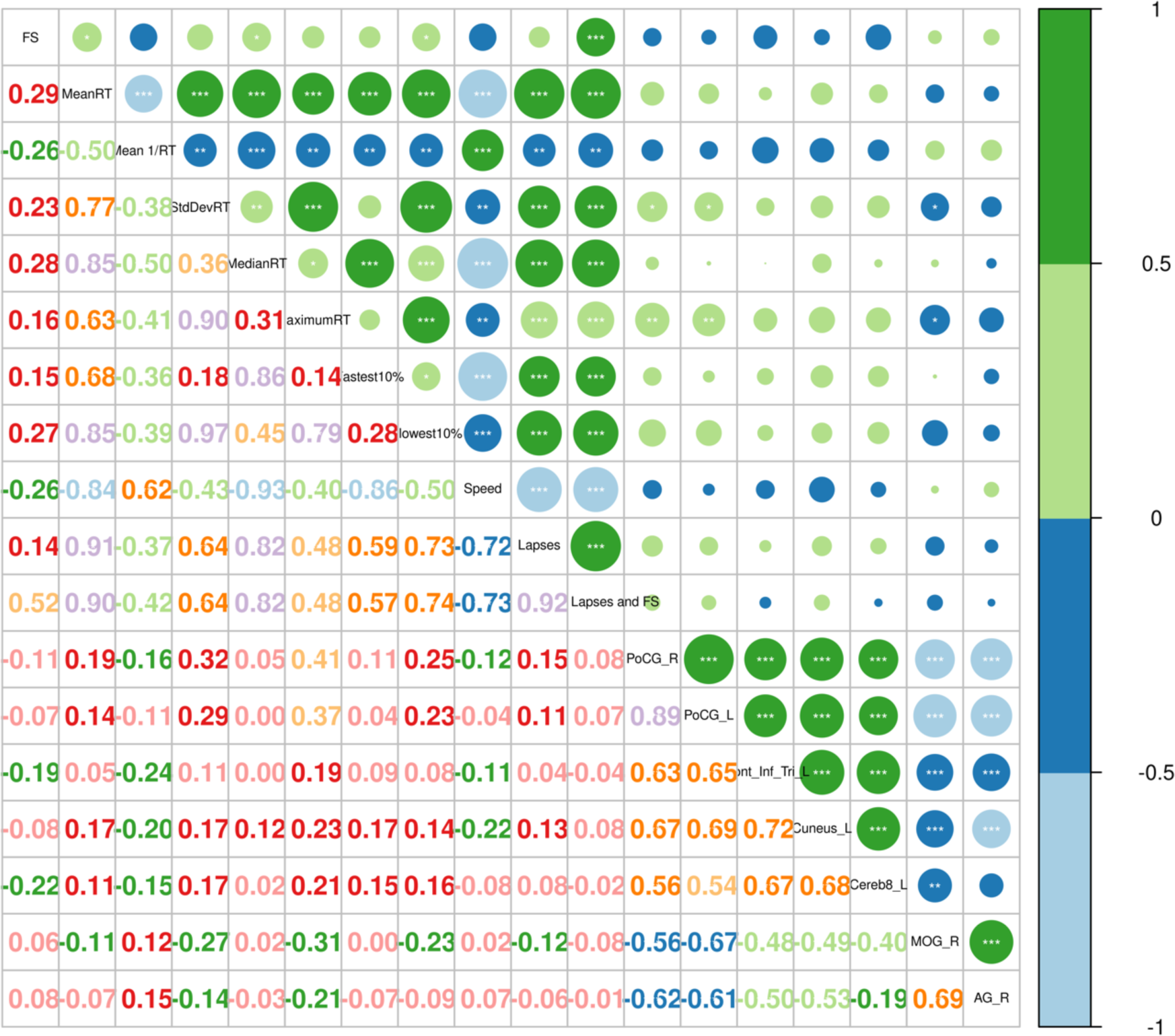
Correlation analysis between vAI_L & PVT

**S3. Spearman.**
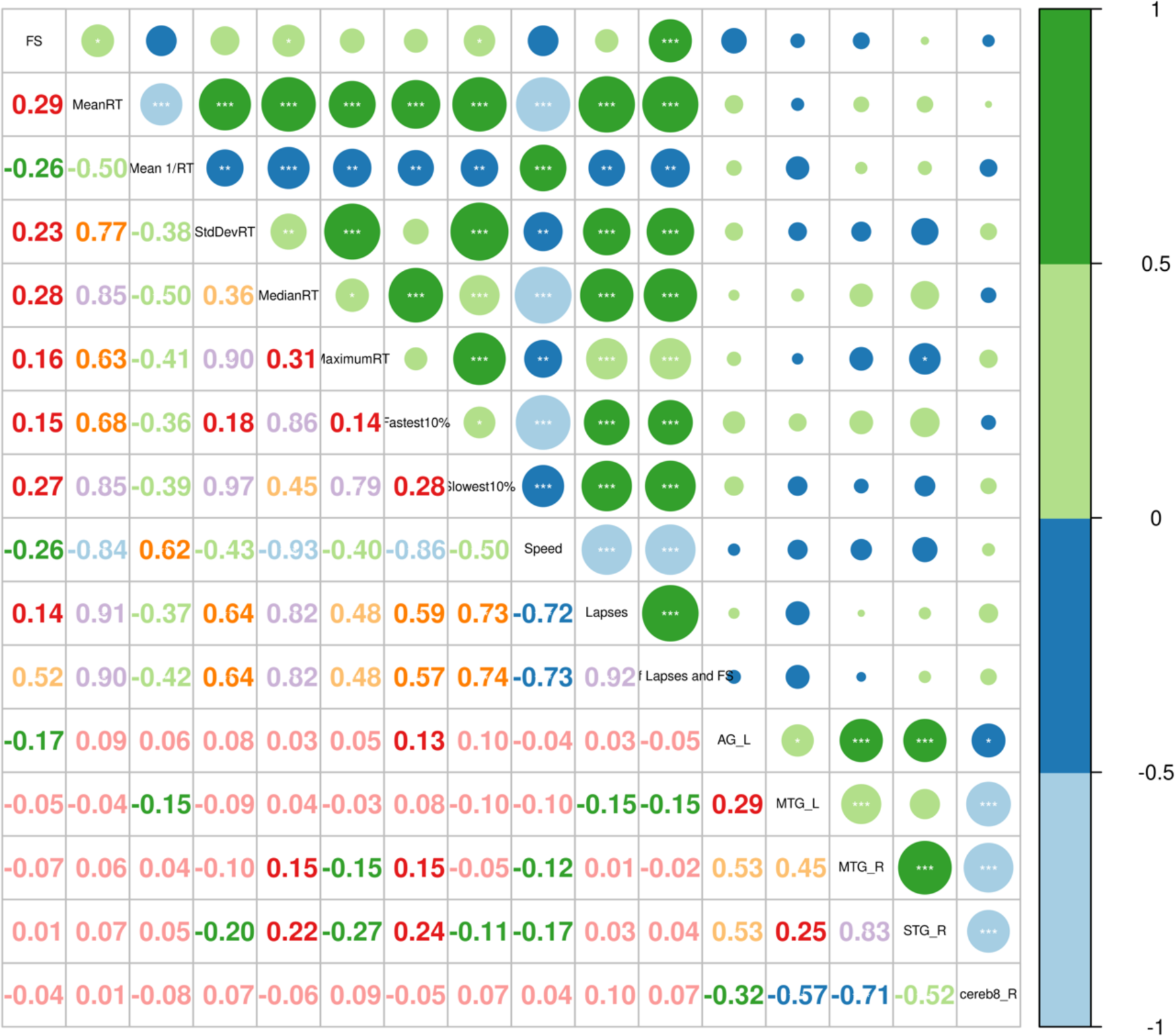
Correlation analysis between dAI_R & PVT

**S4. Spearman.**
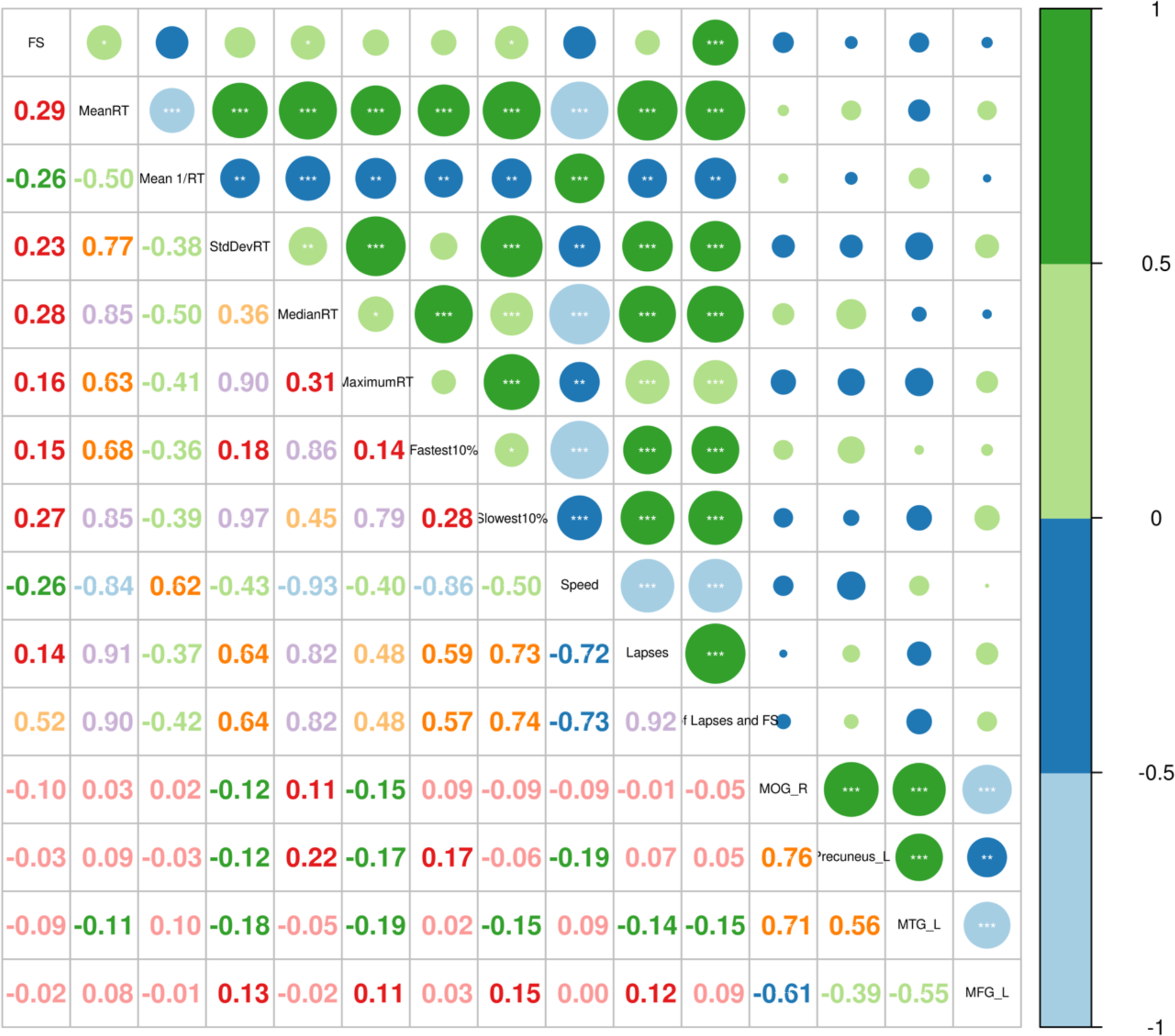
Correlation analysis between dAI_L & PVT

**S5. Spearman.**
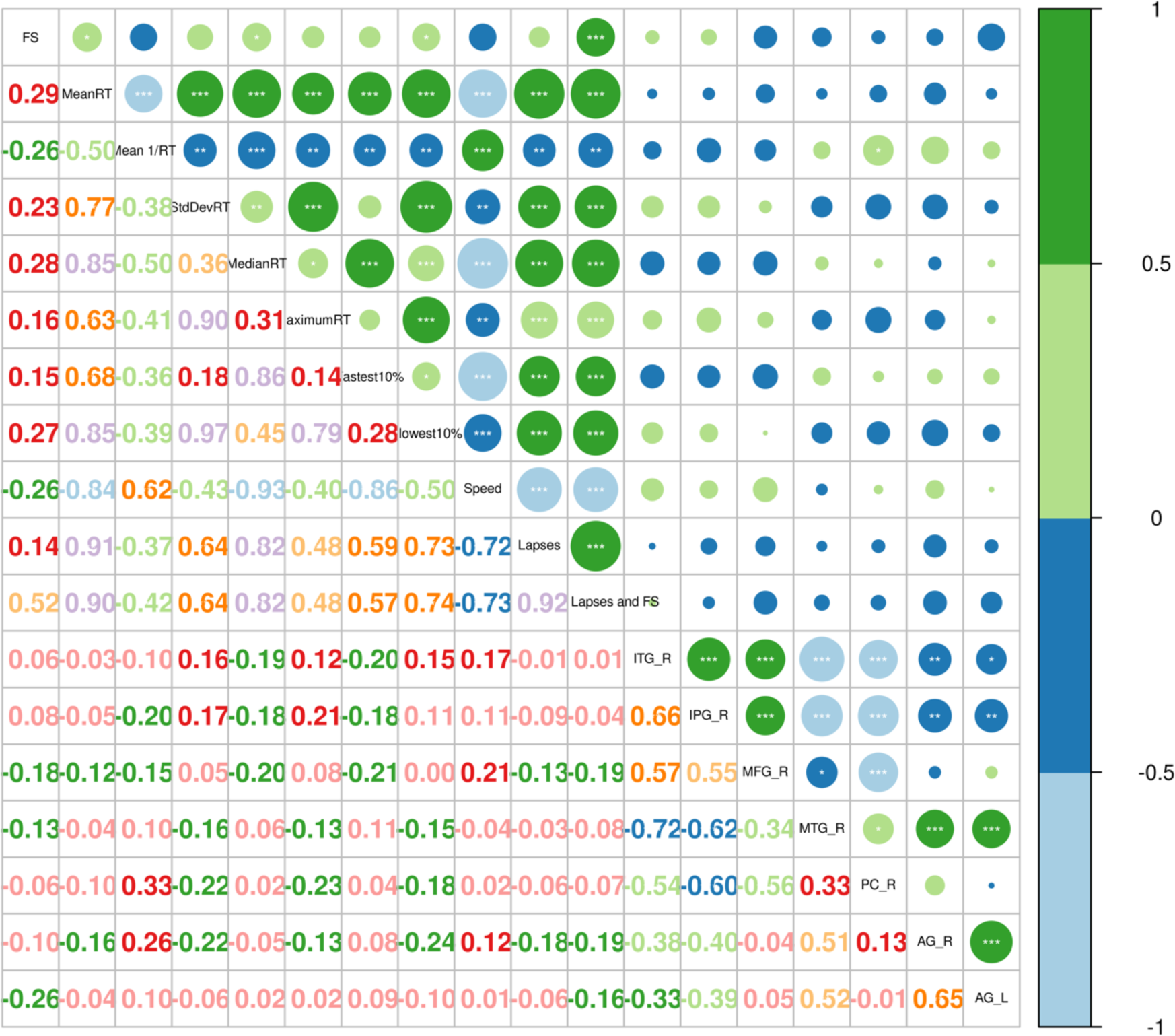
Correlation analysis between PI_R & PVT

**S6. Spearman.**
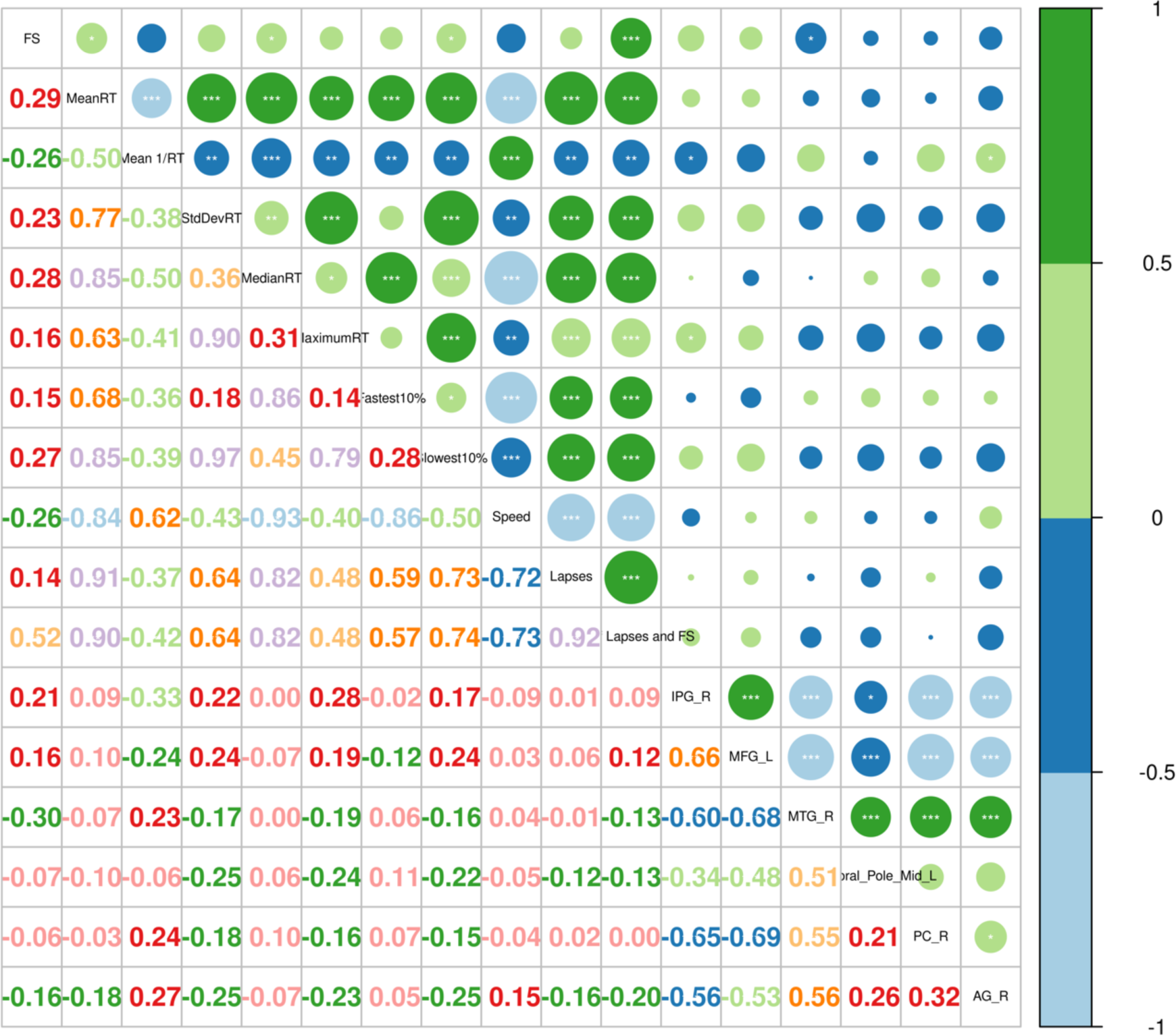
Correlation analysis between PI_L & PVT

## Notes

### Competing Interest Statement

The authors have declared no competing interest.

### Clinical Trial

BM20180040

### Funding Statement

National Key R&D Plan (2022YFC3600500), National Natural Science Foundation of China (No. 82073833 and 8216050478)

### Author Declarations

Institutional Review Board of Beihang University

## References

1. Lowe CJ, Safati A, Hall PA: The neurocognitive consequences of sleep restriction: A meta-analytic review. Neuroscience and biobehavioral reviews 2017, 80:586–604.

2. Lo JC, Ong JL, Leong RL, Gooley JJ, Chee MW: Cognitive Performance, Sleepiness, and Mood in Partially Sleep Deprived Adolescents: The Need for Sleep Study. Sleep 2016, 39(3):687–698.

3. Mason GM, Lokhandwala S, Riggins T, Spencer RMC: Sleep and human cognitive development. Sleep medicine reviews 2021, 57:101472.

4. Goel N, Rao H, Durmer JS, Dinges DF: Neurocognitive consequences of sleep deprivation. Seminars in neurology 2009, 29(4):320–339.

5. Gujar N, Yoo SS, Hu P, Walker MP: The unrested resting brain: sleep deprivation alters activity within the default-mode network. Journal of cognitive neuroscience 2010, 22(8):1637–1648.

6. Gordon EM, Laumann TO, Marek S, Raut RV, Gratton C, Newbold DJ, Greene DJ, Coalson RS, Snyder AZ, Schlaggar BL et al: Default-mode network streams for coupling to language and control systems. Proceedings of the National Academy of Sciences of the United States of America 2020, 117(29):17308–17319.

7. Brandman T, Malach R, Simony E: The surprising role of the default mode network in naturalistic perception. Communications biology 2021, 4(1):79.

8. Smallwood J, Bernhardt BC, Leech R, Bzdok D, Jefferies E, Margulies DS: The default mode network in cognition: a topographical perspective. Nature reviews Neuroscience 2021, 22(8):503–513.

9. Satpute AB, Lindquist KA: The Default Mode Network’s Role in Discrete Emotion. Trends in cognitive sciences 2019, 23(10):851–864.

10. Martin-Subero M, Fuentes-Claramonte P, Salgado-Pineda P, Salavert J, Arevalo A, Bosque C, Sarri C, Guerrero-Pedraza A, Santo-Angles A, Capdevila A et al: Autobiographical memory and default mode network function in schizophrenia: an fMRI study. Psychological medicine 2021, 51(1):121–128.

11. Kaefer K, Stella F, McNaughton BL, Battaglia FP: Replay, the default mode network and the cascaded memory systems model. Nature reviews Neuroscience 2022, 23(10):628–640.

12. Schmahmann JD: The cerebellum and cognition. Neuroscience letters 2019, 688:62-75.

13. Qi J, Li BZ, Zhang Y, Pan B, Gao YH, Zhan H, Liu Y, Shao YC, Zhang X: Altered insula-prefrontal functional connectivity correlates to decreased vigilant attention after total sleep deprivation. Sleep medicine 2021, 84:187–194.

14. Fu W, Dai C, Chen J, Wang L, Song T, Peng Z, Xu M, Xu L, Tang Y, Shao Y: Altered insular functional connectivity correlates to impaired vigilant attention after sleep deprivation: A resting-state functional magnetic resonance imaging study. Frontiers in neuroscience 2022, 16:889009.

15. Goltz D, Pleger B, Thiel SD, Villringer A, Müller MM: Sustained spatial attention to vibrotactile stimulation in the flutter range: relevant brain regions and their interaction. PloS one 2013, 8(12):e84196.

16. Cera N, Castelhano J, Oliveira C, Carvalho J, Quinta Gomes AL, Peixoto MM, Pereira R, Janssen E, Castelo-Branco M, Nobre P: The role of anterior and posterior insula in male genital response and in visual attention: an exploratory multimodal fMRI study. Scientific reports 2020, 10(1):18463.

17. Hopkins WD: Neuroanatomical asymmetries in nonhuman primates in the homologs to Broca’s and Wernicke’s areas: a mini-review. Emerging topics in life sciences 2022, 6(3):271–284.

18. Lo JC, Chong PL, Ganesan S, Leong RL, Chee MW: Sleep deprivation increases formation of false memory. Journal of sleep research 2016, 25(6):673–682.

19. Dinges DF, Orne MT, Whitehouse WG, Orne EC: Temporal placement of a nap for alertness: contributions of circadian phase and prior wakefulness. Sleep 1987, 10(4):313–329.

20. Basner M, Dinges DF: Maximizing sensitivity of the psychomotor vigilance test (PVT) to sleep loss. Sleep 2011, 34(5):581–591.

21. Yan CG, Wang XD, Zuo XN, Zang YF: DPABI: Data Processing & Analysis for (Resting-State) Brain Imaging. Neuroinformatics 2016, 14(3):339–351.

22. Fathy YY, Hepp DH, de Jong FJ, Geurts JJG, Foncke EMJ, Berendse HW, van de Berg WDJ, Schoonheim MM: Anterior insular network disconnection and cognitive impairment in Parkinson’s disease. NeuroImage Clinical 2020, 28:102364.

23. Caligiore D, Mirino P: How the Cerebellum and Prefrontal Cortex Cooperate During Trace Eyeblinking Conditioning. International journal of neural systems 2020, 30(8):2050041.

24. Maldonado T, Bernard JA: The Polarity-Specific Nature of Single-Session High-definition Transcranial Direct Current Stimulation to the Cerebellum and Prefrontal Cortex on Motor and Non-motor Task Performance. Cerebellum (London, England) 2021, 20(4):569–583.

25. Glickstein M, Doron K: Cerebellum: connections and functions. Cerebellum (London, England) 2008, 7(4):589–594.

26. Miller EK: The prefrontal cortex and cognitive control. Nature reviews Neuroscience 2000, 1(1):59-65.

27. Le Merre P, Ährlund-Richter S, Carlén M: The mouse prefrontal cortex: Unity in diversity. Neuron 2021, 109(12):1925–1944.

28. Tanzer M, Derome M, Morosan L, Salaminios G, Debbané M: Cortical thickness of the insula and prefrontal cortex relates to externalizing behavior: Cross-sectional and prospective findings. Development and psychopathology 2021, 33(4):1437–1447.

29. Gamond L, Ferrari C, La Rocca S, Cattaneo Z: Dorsomedial prefrontal cortex and cerebellar contribution to in-group attitudes: a transcranial magnetic stimulation study. The European journal of neuroscience 2017, 45(7):932–939.

30. Diamond A: Close interrelation of motor development and cognitive development and of the cerebellum and prefrontal cortex. Child development 2000, 71(1):44–56.

31. Corbetta M, Shulman GL: Control of goal-directed and stimulus-driven attention in the brain. Nature reviews Neuroscience 2002, 3(3):201–215.

32. Corbetta M, Patel G, Shulman GL: The reorienting system of the human brain: from environment to theory of mind. Neuron 2008, 58(3):306–324.

33. Chica AB, Bartolomeo P, Lupiáñez J: Two cognitive and neural systems for endogenous and exogenous spatial attention. Behavioural brain research 2013, 237:107–123.

34. Japee S, Holiday K, Satyshur MD, Mukai I, Ungerleider LG: A role of right middle frontal gyrus in reorienting of attention: a case study. Frontiers in systems neuroscience 2015, 9:23.

35. Thiel CM, Zilles K, Fink GR: Cerebral correlates of alerting, orienting and reorienting of visuospatial attention: an event-related fMRI study. NeuroImage 2004, 21(1):318–328.

36. Zhang B, Qi S, Liu S, Liu X, Wei X, Ming D: Altered spontaneous neural activity in the precuneus, middle and superior frontal gyri, and hippocampus in college students with subclinical depression. BMC psychiatry 2021, 21(1):280.

37. Li W, Li Y, Yang W, Zhang Q, Wei D, Li W, Hitchman G, Qiu J: Brain structures and functional connectivity associated with individual differences in Internet tendency in healthy young adults. Neuropsychologia 2015, 70:134–144.

38. Tafazoli S, O’Neill J, Bejjani A, Ly R, Salamon N, McCracken JT, Alger JR, Levitt JG: 1H MRSI of middle frontal gyrus in pediatric ADHD. Journal of psychiatric research 2013, 47(4):505–512.

39. Adamaszek M, D’Agata F, Ferrucci R, Habas C, Keulen S, Kirkby KC, Leggio M, Mariën P, Molinari M, Moulton E et al: Consensus Paper: Cerebellum and Emotion. Cerebellum (London, England) 2017, 16(2):552–576.

40. Schutter D: The Cerebellum and Disorders of Emotion. Advances in experimental medicine and biology 2022, 1378:273–283.

41. Koziol LF, Budding D, Andreasen N, D’Arrigo S, Bulgheroni S, Imamizu H, Ito M, Manto M, Marvel C, Parker K et al: Consensus paper: the cerebellum’s role in movement and cognition. Cerebellum (London, England) 2014, 13(1):151–177.

42. Amidfar M, Ko YH, Kim YK: Neuromodulation and Cognitive Control of Emotion. Advances in experimental medicine and biology 2019, 1192:545–564.

43. Cheng CM, Li CT, Tsai SJ: Current Updates on Newer Forms of Transcranial Magnetic Stimulation in Major Depression. Advances in experimental medicine and biology 2021, 1305:333–349.

44. Zhang A, Li G, Yang C, Liu P, Wang Y, Kang L, Wang Y, Zhang K: Alterations of amplitude of low-frequency fluctuation in treatment-resistant versus non-treatment-resistant depression patients. Neuropsychiatric disease and treatment 2019, 15:2119–2128.

45. Sun J, Ma Y, Chen L, Wang Z, Guo C, Luo Y, Gao D, Li X, Xu K, Hong Y et al: Altered Brain Function in Treatment-Resistant and Non-treatment-resistant Depression Patients: A Resting-State Functional Magnetic Resonance Imaging Study. Frontiers in psychiatry 2022, 13:904139.

46. Ameis SH, Fan J, Rockel C, Soorya L, Wang AT, Anagnostou E: Altered cingulum bundle microstructure in autism spectrum disorder. Acta neuropsychiatrica 2013, 25(5):275–282.

47. Kurumaji A, Itasaka M, Uezato A, Takiguchi K, Jitoku D, Hobo M, Nishikawa T: A distinctive abnormality of diffusion tensor imaging parameters in the fornix of patients with bipolar II disorder. Psychiatry research Neuroimaging 2017, 266:66–72.

48. Koshiyama D, Fukunaga M, Okada N, Morita K, Nemoto K, Usui K, Yamamori H, Yasuda Y, Fujimoto M, Kudo N et al: White matter microstructural alterations across four major psychiatric disorders: mega-analysis study in 2937 individuals. Molecular psychiatry 2020, 25(4):883–895.

49. Lee J, Ku J, Han K, Park J, Lee H, Kim KR, Lee E, Husain M, Yoon KJ, Kim IY et al: rTMS over bilateral inferior parietal cortex induces decrement of spatial sustained attention. Frontiers in human neuroscience 2013, 7:26.

50. Luo C, Guo ZW, Lai YX, Liao W, Liu Q, Kendrick KM, Yao DZ, Li H: Musical training induces functional plasticity in perceptual and motor networks: insights from resting-state FMRI. PloS one 2012, 7(5):e36568.

51. Niu M, Rapan L, Funck T, Froudist-Walsh S, Zhao L, Zilles K, Palomero-Gallagher N: Organization of the macaque monkey inferior parietal lobule based on multimodal receptor architectonics. NeuroImage 2021, 231:117843.

52. Torrey EF: Schizophrenia and the inferior parietal lobule. Schizophrenia research 2007, 97(1-3):215–225.

53. Desmurget M, Sirigu A: Conscious motor intention emerges in the inferior parietal lobule. Current opinion in neurobiology 2012, 22(6):1004–1011.

54. Tian Y, Li D, Wang D, Zhu T, Xia M, Jiang W: Decreased Hemodynamic Responses in Left Parietal Lobule and Left Inferior Parietal Lobule in Older Adults with Mild Cognitive Impairment: A Near-Infrared Spectroscopy Study. Journal of Alzheimer’s disease : JAD 2022, 90(3):1163–1175.

55. Anderson AJ, Ren P, Baran TM, Zhang Z, Lin F: Insula and putamen centered functional connectivity networks reflect healthy agers’ subjective experience of cognitive fatigue in multiple tasks. Cortex; a journal devoted to the study of the nervous system and behavior 2019, 119:428–440.

56. Talati A, van Dijk MT, Pan L, Hao X, Wang Z, Gameroff M, Dong Z, Kayser J, Shankman S, Wickramaratne PJ et al: Putamen Structure and Function in Familial Risk for Depression: A Multimodal Imaging Study. Biological psychiatry 2022, 92(12):932–941.

57. Blood AJ, Waugh JL, Münte TF, Heldmann M, Domingo A, Klein C, Breiter HC, Lee LV, Rosales RL, Brüggemann N: Increased insula-putamen connectivity in X-linked dystonia-parkinsonism. NeuroImage Clinical 2018, 17:835–846.

58. Mahone EM, Crocetti D, Tochen L, Kline T, Mostofsky SH, Singer HS: Anomalous Putamen Volume in Children With Complex Motor Stereotypies. Pediatric neurology 2016, 65:59–63.

59. Xu XM, Jiao Y, Tang TY, Zhang J, Salvi R, Teng GJ: Inefficient Involvement of Insula in Sensorineural Hearing Loss. Frontiers in neuroscience 2019, 13:133.

60. Ariani G, Pruszynski JA, Diedrichsen J: Motor planning brings human primary somatosensory cortex into action-specific preparatory states. eLife 2022, 11.

61. Shipp S: The importance of being agranular: a comparative account of visual and motor cortex. Philosophical transactions of the Royal Society of London Series B, Biological sciences 2005, 360(1456):797–814.

62. Palejwala AH, Dadario NB, Young IM, O’Connor K, Briggs RG, Conner AK, O’Donoghue DL, Sughrue ME: Anatomy and White Matter Connections of the Lingual Gyrus and Cuneus. World neurosurgery 2021, 151:e426–e437.

63. Gorgoni M, Sarasso S, Moroni F, Sartori I, Ferrara M, Nobili L, De Gennaro L: The distinctive sleep pattern of the human calcarine cortex: a stereo-electroencephalographic study. Sleep 2021, 44(7).

64. Speed A, Haider B: Probing mechanisms of visual spatial attention in mice. Trends in neurosciences 2021, 44(10):822–836.

65. Chen YC, Spence C: Crossmodal semantic priming by naturalistic sounds and spoken words enhances visual sensitivity. Journal of experimental psychology Human perception and performance 2011, 37(5):1554–1568.

66. Raven F, Van der Zee EA, Meerlo P, Havekes R: The role of sleep in regulating structural plasticity and synaptic strength: Implications for memory and cognitive function. Sleep medicine reviews 2018, 39:3–11.

67. Cavanna AE, Trimble MR: The precuneus: a review of its functional anatomy and behavioural correlates. Brain : a journal of neurology 2006, 129(Pt 3):564–583.

68. Ramage AE, Ray KL, Franz HM, Tate DF, Lewis JD, Robin DA: Cingulo-Opercular and Frontoparietal Network Control of Effort and Fatigue in Mild Traumatic Brain Injury. Frontiers in human neuroscience 2021, 15:788091.

69. Luo Y, Qiao M, Liang Y, Chen C, Zeng L, Wang L, Wu W: Functional Brain Connectivity in Mild Cognitive Impairment With Sleep Disorders: A Study Based on Resting-State Functional Magnetic Resonance Imaging. Frontiers in aging neuroscience 2022, 14:812664.

70. Onitsuka T, Shenton ME, Salisbury DF, Dickey CC, Kasai K, Toner SK, Frumin M, Kikinis R, Jolesz FA, McCarley RW: Middle and inferior temporal gyrus gray matter volume abnormalities in chronic schizophrenia: an MRI study. The American journal of psychiatry 2004, 161(9):1603–1611.

71. Dong QY, Li TR, Jiang XY, Wang XN, Han Y, Jiang JH: Glucose metabolism in the right middle temporal gyrus could be a potential biomarker for subjective cognitive decline: a study of a Han population. Alzheimer’s research & therapy 2021, 13(1):74.

72. Song L, Meng J, Liu Q, Huo T, Zhu X, Li Y, Ren Z, Wang X, Qiu J: Polygenic Score of Subjective Well-Being Is Associated with the Brain Morphology in Superior Temporal Gyrus and Insula. Neuroscience 2019, 414:210–218.

73. Winkelbeiner S, Cavelti M, Federspiel A, Kunzelmann K, Dierks T, Strik W, Horn H, Homan P: Decreased blood flow in the right insula and middle temporal gyrus predicts negative formal thought disorder in schizophrenia. Schizophrenia research 2018, 201:432–434.

74. Briggs RG, Tanglay O, Dadario NB, Young IM, Fonseka RD, Hormovas J, Dhanaraj V, Lin YH, Kim SJ, Bouvette A et al: The Unique Fiber Anatomy of Middle Temporal Gyrus Default Mode Connectivity. Operative neurosurgery (Hagerstown, Md) 2021, 21(1):E8–e14.

75. Pinsk MA, DeSimone K, Moore T, Gross CG, Kastner S: Representations of faces and body parts in macaque temporal cortex: a functional MRI study. Proceedings of the National Academy of Sciences of the United States of America 2005, 102(19):6996–7001.

76. Vogels R: More Than the Face: Representations of Bodies in the Inferior Temporal Cortex. Annual review of vision science 2022, 8:383–405.

77. Studer B, Cen D, Walsh V: The angular gyrus and visuospatial attention in decision-making under risk. NeuroImage 2014, 103:75–80.

78. Rushworth MF, Taylor PC: TMS in the parietal cortex: updating representations for attention and action. Neuropsychologia 2006, 44(13):2700–2716.

79. Humphreys GF, Lambon Ralph MA, Simons JS: A Unifying Account of Angular Gyrus Contributions to Episodic and Semantic Cognition. Trends in neurosciences 2021, 44(6):452–463.

80. Seghier ML: The angular gyrus: multiple functions and multiple subdivisions. The Neuroscientist : a review journal bringing neurobiology, neurology and psychiatry 2013, 19(1):43–61.

81. Ji T, Li X, Chen J, Ren X, Mei L, Qiu Y, Zhang J, Wang S, Xu Z, Li H et al: Brain function in children with obstructive sleep apnea: a resting-state fMRI study. Sleep 2021, 44(8).

82. Delano-Wood L, Stricker NH, Sorg SF, Nation DA, Jak AJ, Woods SP, Libon DJ, Delis DC, Frank LR, Bondi MW: Posterior cingulum white matter disruption and its associations with verbal memory and stroke risk in mild cognitive impairment. Journal of Alzheimer’s disease : JAD 2012, 29(3):589–603.

83. Nakata Y, Sato N, Nemoto K, Abe O, Shikakura S, Arima K, Furuta N, Uno M, Hirai S, Masutani Y et al: Diffusion abnormality in the posterior cingulum and hippocampal volume: correlation with disease progression in Alzheimer’s disease. Magnetic resonance imaging 2009, 27(3):347–354.

